# Microbial contamination of powered air purifying respirators (PAPR) used during the COVID-19 pandemic: an in situ microbiological study

**DOI:** 10.1101/2020.07.30.20165423

**Authors:** Abhijoy Chakladar, Claire G Jones, Jimmy Siu, Mohammed Osman Hassan-Ibrahim, Mansoor Khan

## Abstract

**OBJECTIVE:** To determine whether internal components of powered air purifying respirators (PAPR) used during the Corona virus 2019 disease (COVID-19) pandemic are contaminated with bacteria, fungi and/or any viral material.

**DESIGN & SETTING:** In situ microbiological study. Single NHS Trust, UK.

**OUTCOME MEASURES:** Growth of any bacteria or fungi, or positive polymerase chain reaction results for common respiratory viruses and severe acute respiratory syndrome coronavirus-2 (SARS-CoV-2)

**RESULTS:** 25 PAPR hoods were swabbed; ten (40%) returned positive results. Bacterial growth was detected on six hoods (bacillus simplex, kocuria rhizophilia, bacillus weihenstephensis, microcccus luteus and staphylococcus epidermidis); five of the hoods were positive for fungal growth (non-sporulating environmental mould, NSEM); all sampled hoods tested negative for both SARS-CoV-2 and an expanded panel of respiratory viruses. There was wide variation in the storage of cleaned hoods.

**CONCLUSION:** Despite following recommended cleaning procedures, bacteria and fungi can remain on the internal components of PAPR hoods, at levels significant enough to be swabbed and cultured. PAPR hoods have the potential to cross-infect wearers and patients and are used primarily by clinicians who fail to fit disposable FFP3 respirators; the female sex and non-Caucasian people are less likely to fit FFP3 respirators. The hoods tested cannot be adequately cleaned for use in high risk healthcare environments, PAPR hoods and tubes can act as fomites, and there are evident shortcomings in their provision.

**SUMMARY:** *WHAT IS ALREADY KNOWN ON THIS TOPIC:* Prior to this study there had been no similar investigations looking at the microbiological contamination of PAPR hoods in situ in the real work place hospital environment. SARS-CoV-2 is able to persist in an infective state on surfaces for up to 72 hours. The possibility of internal contamination of PAPR had never been addressed by manufacturers as the hoods were not designed for shared usage.

*WHAT THIS PAPER ADDS:* PAPR hoods in their current form risk transmitting infection between users, and do not protect patients from infected healthcare workers It is possible to detect biological agents within the PAPR hoods despite cleaning as per guidelines An infected asymptomatic carrier is at risk of infecting those around them due to contaminated exhalation

## Introduction

In response to the corona virus 2019 disease (COVID-19) pandemic, NHS and Public Health England have recommended that Health Care Workers (HCW’s) wear a filtering face piece class 3 (FFP3) respirator as part of personal protective equipment (PPE) whenever there is risk of aerosol generating procedures (AGP), and at all times in intensive care units (ICU) or high dependency units (HDU) where COVID-19 patients are treated.^1^

Powered air purifying respirator (PAPR) assemblies, also referred to as power hoods, are used by HCWs requiring high level PPE who either fail a fit test for a disposable FFP3 respirator or for whom prolonged use of a standard FFP3 respirator is intolerable. These are utilised during the highest risk aerosol generating procedures such as tracheostomy insertion and have been shown to offer better conditions for the wearer when performing complex medical procedures versus reusable respirators.^2,3^ There is evidence to suggest that females and non-Caucasian people are less likely to pass fit tests for FFP3 respirators.^4-9^

There are currently approximately 100 PAPRs in our Trust; PAPR assemblies consist of a fan unit with battery pack and filters, connected via corrugated tubing to a headtop or hood (figure 1). The majority of hoods are *3M Scott Safety FH1 headtops*, the rest are *3M Scott Safety FH2 PAPR headtops* (3M Scott, Skelmersdale, UK). The *3M Scott Safety FH1 headtop* is a half hood that provides a loose seal around the face; the *3M Scott Safety FH2 PAPR headtop* is the similar to the *FH1* but, in addition, has a shoulder cape that provides neck and shoulder protection. The filtered air, under positive pressure is supplied to the wearer; this and the wearer’s exhaled breath is expelled around the edges of the hood and through an unshrouded expiratory valve at the bottom of the visor component of the hood. The hoods consist of a clear polyethylene terephthalate glycol visor, stitched to fabric, with a neoprene margin that sits against the wearers face. These PAPR and hoods were originally developed to provide respiratory protection in industrial use to help combat pulmonary contamination by inanimate particulates; their use has been adapted for the current circumstances during the COVID-19 pandemic by upgrading the filters attached to the fan unit.

**Figure 1.**
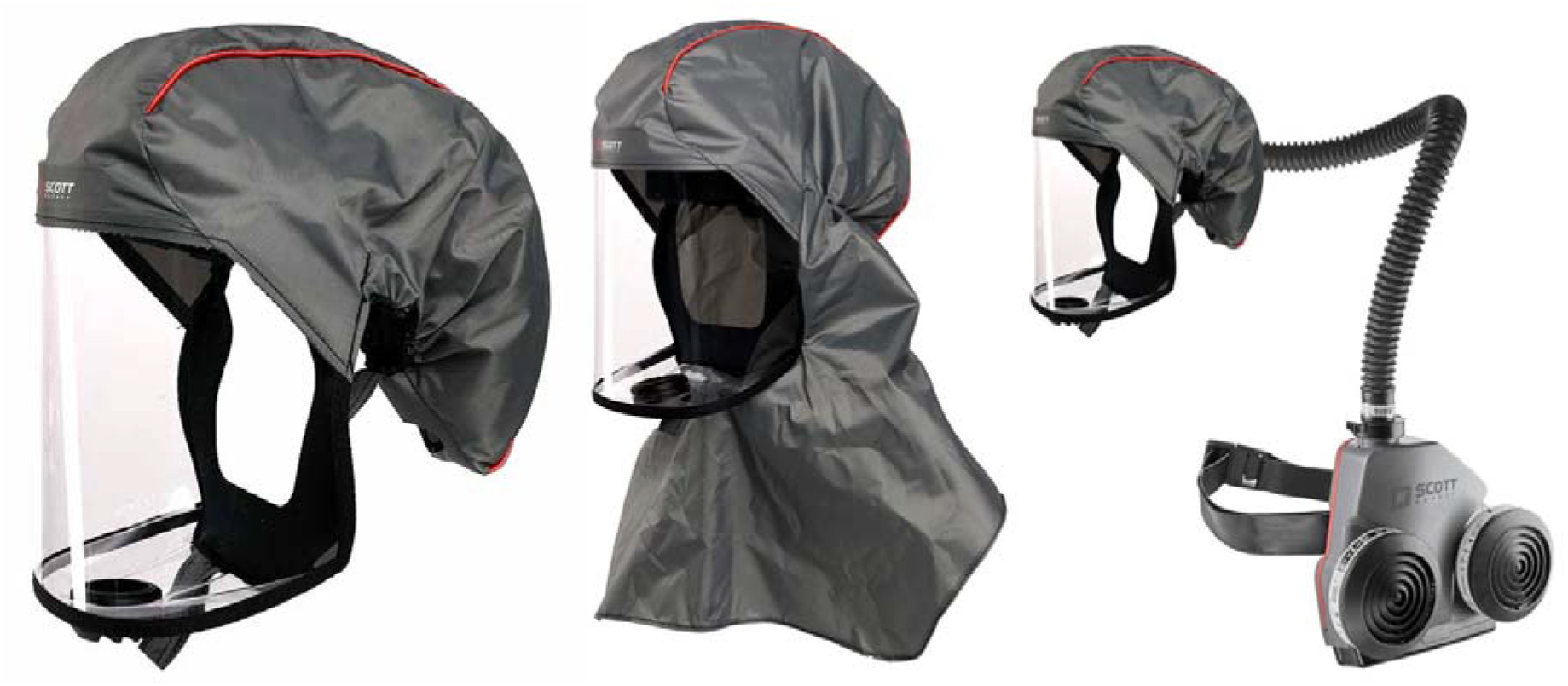
3M Scott FH1 and FH2 *Headtops* and PAPR arrangement

The fidelity of the hood relies on the assumption that high flow forced air prevents the wearer from inhaling directly from the environment. This is also the rationale behind the supposition that users are unable to contaminate the internal spaces of the hood such as the tubing because it is felt the high air flow prevents any retrograde flow of exhaled air. Local guidance states that users should don the hood before turning on the fan unit. In addition, obstruction of the filter air inlet ceases airflow to the hood. This is of particular significance in the current pandemic since it is now well documented that individuals may be COVID-19 positive but asymptomatic,^10^ thus putting other shared hood users at risk.

Particular concerns have been expressed by HCWs regarding the infection control of PAPRs: the potential to become fomites via handling; their contamination by aerosol deposition of respiratory secretions, or other transmission routes;^11^ and lack of confidence in the cleaning and decontamination process. Current local guidelines from infection control services recommend the use of Clinell Universal Wipes (benzalkonium chloride 0.54g, didecyldimonium chloride 0.54g, and phenoxyethanol 0.6g per 100g; GAMA Healthcare, Watford, UK) for cleaning and disinfection but does not address the cleaning of the internal tubing of the power hoods.^12^ There is little research available examining the efficacy of different cleaning methods, but both cleaning alone and cleaning plus disinfection have been shown to be effective in removing the influenza virus from PAPR.^11^

PAPR hoods are intended for single user use and as a result any potential contamination of the internal components of the hood has not been addressed by the manufacturer and nor does it form part of any standard decontamination protocols.^12^ Evidence suggests that viral particles may persist on surfaces, in an infectious state, for up to 72 hours,^13^ and there is concern that PAPRs may potentially contaminate sterile fields.^14^ We were concerned that PAPR hoods may pose an infection risk between users as internal parts of the respirators, for example, the corrugated breathing tube, cannot be cleaned effectively using standard procedures.

We conducted a study to determine whether a sample of the hood and tubing components of PAPR assemblies used at our institution were colonised with bacteria, fungi and whether any viral DNA or RNA could be detected. This study does not address the additional concern that exhalation around the PAPR hood and via unshrouded, unfiltered, expiratory valves does not protect patients from potential infection by infected hoods or their wearers.

## Method

### Study design and setting

After approval from the BSUH NHS Trust Executive Board, we conducted a study to test the PAPR hoods in use in our institution by swab testing them for evidence of bacterial, fungal, common respiratory viruses and severe acute respiratory syndrome coronavirus-2 (SARS-CoV-2) contamination.

### Swabbing

PAPR hoods were tested in the clinical environments in which they were stored and used. Each PAPR hood had four swabs: two charcoal and two viral, one of each to the inside of the visor and the inside of the corrugated tube respectively.

The hood-holder manipulated the mask to allow the swab-taker access to the target area to take samples (Figure 2). Each PAPR was swabbed for bacteria, fungi, respiratory viruses (extended respiratory panel) and SARS-CoV-2. Bacterial and fungal samples were taken with Transwab® Amies Charcoal swabs (MW171, Medical Wire & Equipment, UK), and viral samples taken with Sigma Virocult® swabs (MW951S, Medical Wire & Equipment, UK).

Swabs were moistened with sterile 0-9% sodium chloride solution. Swabs were taken from two pre-defined area: first, inside of the clear plastic visor including the expiratory valve, and second, inside the corrugated tube attached to the hood. The visor and expiratory valve were divided in to two halves, one for the charcoal swab and one for the viral swab. The expiratory valve was swabbed first with care taken to apply the swabs uniformly over the surface and then over the respective side of the visor in linear, up-down, fashion with care taken to access any corners and cover the full area of the visor. The corrugated tubes were also divided in two halves, one for each type of swab. The swab was passed from the rim down to approximately 10cm, the length of the swabs, with care taken to swab within the ridges of the corrugations.

### Investigators and PPE

All samples were taken by consultant anaesthetists experienced in the wearing of FFP3 respirators, PPE and conducting procedures with aseptic non-touch technique (ANTT). Investigators wore PPE to protect themselves from potentially colonised PAPR and to prevent colonisation of the equipment by the investigators. The ‘hood-holder’ wore fresh non-latex sterile gloves, eye protection, and FFP3 respirator; the ‘swab-taker’ wore fresh non-latex sterile gloves, surgical gown, eye protection, and FFP3 respirator; the investigator responsible for sample labelling and bagging wore non-sterile latex free gloves, eye protection, and a FFP3 respirator.

### Traceability and labelling

The location of PAPR across the Trust was identified. Hoods were marked with a unique code that corresponded with the code for their respective swabs and allowed for the location of any PAPR to be identified. For example, ‘RSCH_ENT_1 ’ refers to a PAPR that is kept in the ENT outpatients at the Royal Sussex County Hospital. Where the PAPR already had a number designation in a clinical area, this was reflected in the assigned code.

## Laboratory testing and reporting

### Bacterial testing

Transwab® Amies Charcoal swabs were cultured onto Blood (OXOID Colombia Agar with horse blood, Code: PB0122, distributed by ThermoFisher, UK) and Chocolate (Oxoid Colombia Agar with Chocolated Horse Blood, Code: PB0124, ThermoFisher, UK) agar plates and incubated in carbon dioxide (CO_2_) for 24 hours. Any growth was identified using the MALDI-TOF system (Bruker, USA).

### SARS-CoV-2 (COVID-19) testing

Testing for SARS-CoV-2 (COVID-19) was performed by real-time RT-PCR method. 400 μl of each of the Sigma Virocult® swabs with added 5μl internal control were extracted using the Kingfisher Flex system (ThermoFisher, UK) with elution volume of 50 μl. 10 μl of extracted sample material was then added to 15 μl PCR master mix of the CE-marked Bosphore® Novel Coronavirus (2019-nCoV) Detection v2 Kit (manufactured by Anatolia Geneworks, Turkey and supplied by Launch Diagnostics, UK). This is then amplified on the Applied Biosystems 7500 machine (Applied Biosystems, USA) using the thermal protocol for Bosphore® Novel Coronavirus (2019-nCoV) Detection Kit v2 as per manufacturer recommendation. The targets for the test include (orf1ab & Egene). The manufacturer reported 100% specificity and 95% sensitivity (with analytical detection limit of 25 copies/rxn).

### Respiratory panel testing

Testing for the respiratory panel was performed by real-time RT-PCR method. 400 μl of each of the Sigma Virocult® swabs was extracted using the Nuclisens EasyMag system (Biomerieux, France) with elution volume of 100 μl. The respiratory panel multiplex RT-PCR detection kit (CE-marked Bosphore® Respiratory pathogen panel v6, manufactured by Anatolia Geneworks, Turkey and supplied by Launch Diagnostics, UK) 7 mini-multiplexes with each of the mini-multiplexes has their own integrated internal control. This multiplex RT-PCR panel can detect 19 targets including: influenza A (pandemic H1N1 or non-pandemic H1N1), influenza B, parainfluenza 1-4, RSV, Enterovirus, Parechovirus, Rhinovirus, seasonal Coronaviruses (NL63, 229E, OC43 & HKU), Human metapneumovirus, Human bocavirus, Adenovirus, and Mycoplasma pneumoniae. 10 μl of extracted sample material was then added to 15 μl PCR master mix of each of the mini-multiplexes. These were then amplified on the Applied Biosystems 7500 Fast machine (Applied Biosystems, USA) using the thermal protocol for Bosphore® Respiratory pathogens panel detection kit v6 as per manufacturer recommendation. The manufacturer reported 100% specificity and 95% sensitivity.

### Fungal testing

Transwab® Amies Charcoal swabs were cultured onto Sabouraud dextrose agar plates (Oxoid Sabouraud dextrose agar with Chloramphenicol, Code: PO0161, ThermoFisher, UK) and incubated in air at 35°C for 5 days. Any growth of fungi was then identified macroscopically and/or microscopically as appropriate.

### Patient and public involvement

It was not appropriate or possible to involve patients or the public in the design, or conduct, or reporting, or dissemination plans of our research. This was a microbiological study with no patient or public contact.

## Results

Over a 48 hours period in June 2020, 25 (approximately 25%) PAPR hoods and their attached corrugated tubes were tested for evidence of bacterial, fungal, respiratory viral (extended respiratory panel, and SARS-CoV-2 colonisation, generating a total of 100 swabs for analysis.

Out of the 25 hood and tube assemblies swabbed, ten (40%) returned a positive result (Table 1). See supplementary section 1 for the results from all swabs taken.

### Bacterial testing

There was bacterial growth detected on six PAPR hoods. Bacteria were detected on the visor only of three hoods, corrugated tube only of two PAPR hood. Bacteria were detected on the visor and corrugated tube of one hood. The bacteria detected were bacillus simplex, kocuria rhizophilia, bacillus weihenstephensis, microcccus luteus and staphylococcus epidermidis.

### SARS-CoV-2 (COVID-19) and respiratory panel testing

All PAPR hoods sampled tested negative for SARS-CoV-2 (COVID-19) and respiratory viral panel testing including influenza A (pandemic H1N1 or non-pandemic H1N1), influenza B, parainfluenza 1-4, RSV, Enterovirus, Parechovirus, Rhinovirus, seasonal Coronaviruses (NL63, 229E, OC43 & HKU), Human metapneumovirus, Human bocavirus, Adenovirus, and Mycoplasma pneumoniae.

### Fungal testing

Five of the PAPR hoods were positive for fungal growth; all grew non-sporulating environmental mould (NSEM) which could not be identified.

### Hood storage

We noted a wide variation in the storage of ‘clean’ hoods, ranging from lying open on a trolley next to a ward bay accommodating patients with COVID-19, to uncontrolled storage in boxes, to controlled storage, after double cleaning, tracking locations of use, a strict log of previous wearers, cleaning diaries and storage in the same box.

**Table 1.** Positive swab results.

| # | Hosp | Ward | Hood | Model | Part | Code | Bacteria | Viral panel | SARS-Cov-2 | Fungal |
| --- | --- | --- | --- | --- | --- | --- | --- | --- | --- | --- |
| 4 | PRH | Pyecombe | 1 | FH1 | Visor | PRH_PYE_1_visor | Bacillus simplex | x | x | x |
| 12 | PRH | ICU | 5 | FH1 | Visor | PRH_ICU_5_visor | x | x | x | NSEM |
| 13 | PRH | Theatre | 1 | FH1 | Visor | PRH_Theatre_1_visor | x | x | x | NSEM |
| 15 | RSCH | Labour Ward | 2 | FH1 | Visor | RSCH_L13_2_visor | x | x | x | NSEM |
| 16 | RSCH | ENT OP | 6 | FH1 | Tube | RSCH_ENT_6_tube | Kocuria rhizophilia | x | x | x |
| 19 | RSCH | ENT OP | 4 | FH1 | Visor | RSCH_ENT_4_visor | x | x | x | NSEM |
| 20 | RSCH | ENT ward | 1 | FH1 | Visor | RSCH_L8AW_1_visor | Bacillus weihenstephensis | x | x | x |
| 21 | RSCH | ED | 1 | FH2 | Tube | RSCH_ED_1_tube | Micrococcus luteus | x | x | x |
| 23 | RSCH | ICU | 2 | FH1 | Visor | RSCH_ICU_2_visor | Staphylococcus epidermidis | x | x | NSEM |
| 25 | RSCH | ICU | 4 | FH2 | Visor | RSCH_ICU_4_visor | Micrococcus luteus | x | x | x |
| 25 | RSCH | ICU | 4 | FH2 | Tube | RSCH_ICU_4_tube | Micrococcus luteus | x | x | x |

PRH=Princess Royal Hospital. RSCH=Royal Sussex County Hospital. FH1/FH2=model of 3M Scott PAPR Headtop. ICU=intensive care unit; ED=emergency department; NSEM=non-sporulating environmental mould; x=negative swab; ENT=ear, nose, and throat; OP=outpatient

## Discussion

To our knowledge this is the first study to examine the internal components of PAPR hood assemblies for contamination with a wide range of pathogens including SARS-CoV-2 and shows that despite following recommended cleaning procedures, bacteria and fungi can remain on the internal components of the hoods, at a level significant enough to be swabbed and cultured. Viral material, in particular SARS-CoV-2, was not detected.

Staphylococcus epidermidis, Kocuria rhizophilia, and Microcccus luteus are Gram positive cocci. Staphylococcus epidermidis is a skin coloniser that can cause infections of intravenous lines and prosthetic materials, for example artificial heart valves and orthopaedic joints. Kocuria rhizophiliais found in the oropharynx, oral mucosa, and skin, it does not usually cause of human infections but can rarely can cause opportunistic infection in the severely immunocompromised. Microcccus luteus is part of the normal bacterial flora of human skin and is not generally pathogenic. Bacillus simplex and Bacillus weihenstephensis are gram positive bacilli that can colonise skin and our found in soil; they are unlikely to be pathogens in humans, however Bacillus simplex has been implicated in food borne disease and as a potential cause of brain abscess.^15^ The significance of NSEM is difficult to qualify as they are diverse and, as they do not sporulate, are difficult to identify; they are usually not pathogenic but should be considered in the presence of immunocompromised patients. However, one of the four hoods sampled that benefited from controlled storage, after double cleaning, tracked locations of use, and strict log of previous wearers with a cleaning diary, had not been worn for 6 days, and grew a NSEM.

The significance of risk to wearers is difficult to quantify, but it is reasonable to infer that if the organisms above are able to survive, other, more pathogenic, organisms have the potential to survive placing wearers at risk of cross infection from colleagues. PAPR are used primarily by clinicians who fail to fit disposable FFP3 respirators and those that conduct the highest risk procedures. Contamination of hood assemblies is not surprising given the advice is to don the PAPR hood before turning on the fan unit, and if the air inlet to the filter is blocked, airflow to the hood ceases, allowing exhaled breath to circulated within the hood and up towards the tubing. Confidence in PPE in the UK is low and there are concerns that female healthcare workers and those from Black, Asian and minority ethnic backgrounds are less likely to fit respirators adequately as PPE design is for the most part based on physical characteristics of males from Europe and the USA.^4-9^ Research suggests that the size and shape of wearers’ faces, have an impact on the success of passing fit tests for respirators, with people with wider faces and prominent noses more likely to pass.^16-19^ There is a dearth of comparative studies including people from south and east Asia and this combines with a one size or model fits all approach to respirator provision suggests an element of structural sex and racial bias in the provision of respiratory PPE internationally; our findings compound this.

The charcoal and viral swabs are intended for mucosal and not surface sampling. We dampened the swabs to recreate this but it may have reduced the chances of success of a positive test. It may be that the swabbing techniques utilised are not able to detect viral contamination. We carefully planned the swabbing so that each swab consistently sampled the same area of respective hoods and tubes, minimising overlap and the potential to remove organisms. We considered the risk of contamination by the swab collecting team; however, fresh sterile PPE was donned by the ‘swab taker’ for each hood and ‘hood holder’. The hoods were tested in the order their results are presented and the distribution of organisms would suggest that investigators did not cross-contaminate hoods. Without a suitable log or tracking method, we were unable to ascertain when the hoods had last been worn. It was our impression that all hoods except one had been worn within 24 hours; a longer time length of time since last use may reduce the chance of successfully culturing bacteria and after 72 hours it is difficult to retrieve viral materials from hard surfaces. One hood assembly was last worn 6 days prior to swabbing, and this grew a NSEM. We examined 25 hoods and tube assemblies, approximately 25% of our institutions stock in circulation; we believe this is an adequate sample to test our theory.

During this pandemic, confidence in PPE has been reduced by conflicting advice and difficulties with availability.^20^ Despite the risk to HCWs, when collecting the swabs there was empirical evidence of people who had failed fit testing to a disposable FFP3 respirators, but continued to wear them as they were fearful of the cross infection risk with PAPRs.^21-24^

The PAPR and hood systems are not designed for medical environments and there is concern regarding their efficacy in maintaining a sterile environment.^14^ Simulated microbiological research has demonstrated that PAPR systems may be as effective at fluid resistant surgical masks (FRSM) at maintaining a sterile field in the operating theatre,^21^ however, care must be taken when extrapolating results as all not all PAPR hoods are the same; the hoods in the study had a permeable fabric filter to aid the exhaust of gas, the hoods employed at our institution have an expiratory valve that exhausts unfiltered gas (a mix of exhaled and fresh air) directly in to what would be a sterile surgical field. There is a risk to patients, depending on the design of the PAPR hood, posed by HCWs with infections exhaling unfiltered air directly over them,^14,22^ with gas driven at high flow rates, and from hoods that are colonised by organisms from previous wearers.

The hoods employed in our PAPR assemblies are not designed for clinical use, with a high likelihood of biological contamination. It is difficult to clean fabric and although tubes without foam components can be immersed, hoods cannot be soaked in cleaning fluid, washed in a washing machine or respirator washer, or sterilised with ethylene dioxide, radiation or steam (risk of damage to components) or vaporized hydrogen peroxide (VHP) as they may harmful residual levels of residual VHP;^12^ strong cleaning products, such as disinfectant wipes, may have an adverse effect on the product;^23^ Cleaning should be performed by trained individuals, and not, as is often the case, by healthcare workers after long shifts. There is no evidence based guidance from manufacturers for the disinfection of the internal parts of the PAPR hood assemblies as they are expected to be single user use,^12^ recent studies have shown that PAPRs can be adequately cleaned and disinfected in a laboratory environment, but the PAPRs were from a different manufacturer, and the cleaning and disinfection method employed was more complicated than the locally recommended method.^11,23^ The storage of PAPR assemblies was variable and in some cases inadequate, which may lead to their contamination in between use. With any potential risk of cross infection, it is imperative that if a user falls ill, previous users of any PAPR hood assemblies can be traced effectively. Current storage methods do not allow for this except at a few locations.

### Recommendations/options to mitigate risks posed by PAPRs

1. Stock multiple, at least more than one FFP3 brand or model to improve fit test pass rates
2. Issue reusable respirators to those that fail fit tests or those that regularly perform high risk procedures
3. Provide HCWs with personal PAPR hoods and tubes; battery, fan, belts, and filter packs can be shared. The costs of a *3M Scott Safety FH1/FH2 Headtop* and corrugated tube are £46.29 and £39.64 respectively [quote from distributor]
4. Work with industry to define a definitive and effective cleaning method for PAPR assemblies that will be shared
5. Train staff to safely and effectively clean PAPR assemblies
6. Keep a strict log of recent users and cleaning to enable tracking and tracing of staff that have worn a PAPR hood, in the event that a staff member falls ill from or develops COVID-19
7. The use of PAPR hood assemblies with unshrouded expiratory valves during surgery should be reviewed
8. Consider alternative cleaning methods for hoods and tubes
9. Disposable FFP3 respirator design needs to be improved and steps taken to improve fit and tolerability;^24,25^ FFP3 respirators personalised to the wearers face shape should be considered
10. Collaborate with industry to develop PAPR systems specifically designed for healthcare

## Conclusion

This study demonstrates that current cleaning guidelines are not effective in disinfecting PAPR hood assemblies and any microbes that survive the cleaning process can persist for more than 5 days. PAPR are an important component in the range of respiratory PPE available to healthcare staff and this is unlikely to be the last pandemic.^26-28^ Current UK/PHE PPE guidance is not in line with other international standards.^29^ We have made recommendations regarding respiratory PPE for staff to mitigate the cross infection risk posed to both staff and patients by PAPR hoods and tubes, acting as fomites, and the evident structural sex and racial bias in the provision of respiratory PPE. It is our opinion that, given the constraints outlined by both the manufacturer and the distributor, the hoods tested cannot be adequately cleaned for use in high risk healthcare environments. Adequate PPE, their use, and effective cleaning should be researched urgently, and industry collaboration funded to develop PAPR systems that are fit for purpose in high risk healthcare settings.

## Data Availability

All data is presented in the manuscript

## Acknowledgements

We would like to acknowledge: Dr George Findlay, Managing Director, Chief Medical Officer and Deputy Chief Executive, Brighton and Sussex University Hospitals NHS Trust for his support; Dr Sunil Sharma, Consultant in Microbiology & Infectious Diseases, for his guidance regarding bacterial and fungal growths; and Lucy Francis, Trust Clinical Procurement Manager for her support regarding PAPR purchasing and cleaning guidance.

## Contributors

All authors contributed to study design, data interpretation, and writing of the report. AC, CJ, and JS collected the data. MOH-I oversaw the laboratory tests and reporting, and interpreted the microbiology and virology data. AC is the guarantor. The guarantor accepts full responsibility for the work and/or the conduct of the study, had access to the data, and controlled the decision to publish. All authors approved the final version of the manuscript. The corresponding author attests that all listed authors meet authorship criteria and that no others meeting the criteria have been omitted.

## No competing interests

All authors have completed the ICMJE uniform disclosure form at www.icmje.org/coi_disclosure.pdfand declare: no support from any organisation for the submitted work; no financial relationships with any organisations that might have an interest in the submitted work in the previous three years; no other relationships or activities that could appear to have influenced the submitted work.

## Ethical approval

Not required.

The manuscript’s guarantor (AC) affirms that the manuscript is an honest, accurate, and transparent account of the study being reported; that no important aspects of the study have been omitted; and that any discrepancies from the study as planned (and, if relevant, registered) have been explained.

## Supplementary information

### 1. Swabbing process

1. Identify power hood
2. Label power hood (same name as sample)

- E.g. Pye_PRH_1 (Pyecombe, PRH, Hood 1)
- E.g. ICU_RSCH_3 (ICU, RSCH, Hood 3)
3. Don PPE:

- Hood holder – fresh sterile gloves, eye protection, and FFP3 respirator
- Swab taker – fresh sterile gloves, eye protection, and FFP3 respirator
- Swab labelling & bagger – non-sterile gloves, eye protection, and FFP3 respirator
4. Prepare power hood and hold as per illustration
5. Swabbing

- plastic visor and the exhalation vent

⊳ take BLACK (bacterial/fungal) swab and drip on sterile 0.9% NaCl
- Wipe swab over (inside of) clear face piece and expiration vent

⊳ take GREEN (viral/COVID) swab and drip on sterile 0.9% NaCl
- Wipe swab over (inside of) clear face piece and expiration vent
- Corrugated tube at hood end

⊳ take BLACK (bacterial/fungal) swab and drip on sterile 0.9% NaCl

- Wipe swab around and 10 cm in to corrugated tube

⊳ take GREEN (viral/COVID) swab and drip on sterile 0.9% NaCl

- Wipe swab around and 10 cm in to corrugated tube
6. Labelling samples

- E.g. Pye_PRH_1_Bact/Fung_visor = Pyecombe, PRH, Hood 1, bacterial/fungal swab from visor
- E.g. ICU_RSCH_3 _Viral_tube = ICU, RSCH, Hood 3, viral & COVID swab from corrugated tube
7. Bagging

- Each sample must go in its own labelled bag

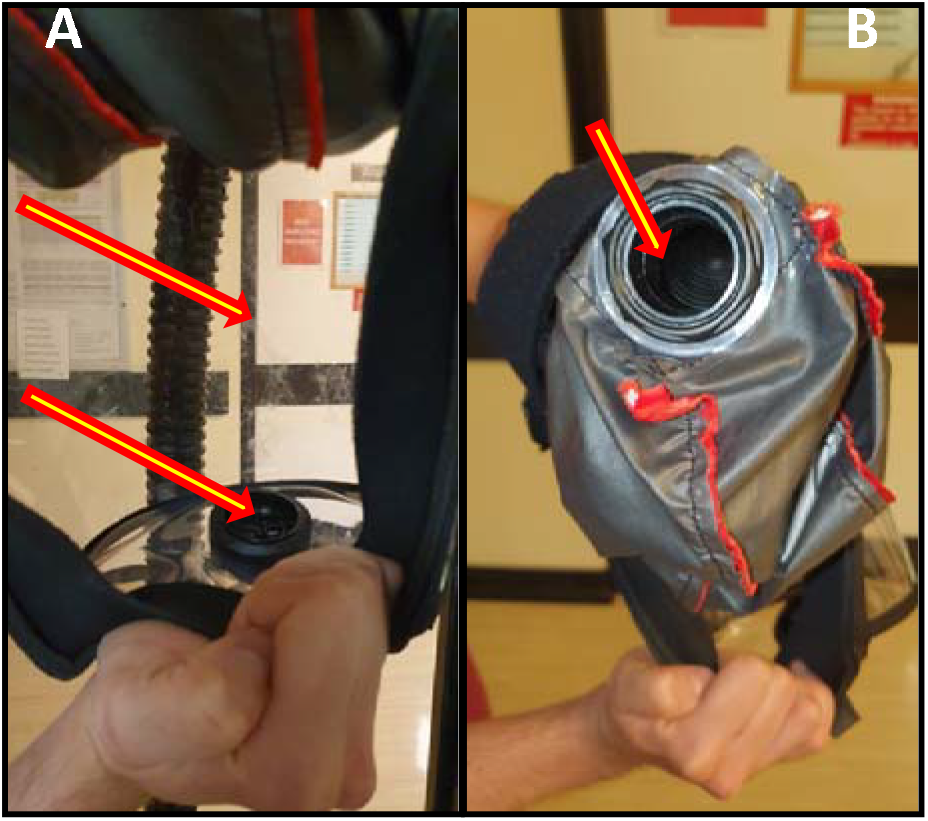

### 2. Complete results table

| # | Hosp | Ward | Hood | Model | Part | Code | Bacteria | Viral panel | SARS_Cov-2 | Fungal | Comments |
| --- | --- | --- | --- | --- | --- | --- | --- | --- | --- | --- | --- |
| 1 | PRH | Labour Wd | 1 | FH1 | Visor | PRH_CDS_1_visor | x | x | x | x |  |
|  |  |  |  |  | Tube | PRH_CDS_1_tube | x | x | x | x |  |
| 2 | PRH | Labour Wd | 2 | FH1 | Visor | PRH_CDS_2_visor | x | x | x | x |  |
|  |  |  |  |  | Tube | PRH_CDS_2_tube | x | x | x | x |  |
| 3 | PRH | Labour Wd | 3 | FH1 | Visor | PRH_CDS_3_visor | x | x | x | x |  |
|  |  |  |  |  | Tube | PRH_CDS_3_tube | x | x | x | x |  |
| 4 | PRH | Pyecombe | 1 | FH1 | Visor | PRH_PYE_1_visor | Bacillus simplex | x | x | x | Dirt on swab |
|  |  |  |  |  | Tube | PRH_PYE_1_tube | x | x | x | x |  |
| 5 | PRH | Pyecombe | 2 | FH1 | Visor | PRH_PYE_2_visor | x | x | x | x |  |
|  |  |  |  |  | Tube | PRH_PYE_2_tube | x | x | x | x |  |
| 6 | PRH | ED | 1 | FH1 | Visor | PRH_ED_1_visor | x | x | x | x |  |
|  |  |  |  |  | Tube | PRH_ED_1_tube | x | x | x | x |  |
| 7 | PRH | ED | 2 | FH1 | Visor | PRH_ED_2_visor | x | x | x | x |  |
|  |  |  |  |  | Tube | PRH_ED_2_tube | x | x | x | x |  |
| 8 | PRH | ICU | 1 | FH2 | Visor | PRH_ICU_1_visor | x | x | x | x |  |
|  |  |  |  |  | Tube | PRH_ICU_1_tube | x | x | x | x |  |
| 9 | PRH | ICU | 2 | FH1 | Visor | PRH_ICU_2_visor | x | x | x | x |  |
|  |  |  |  |  | Tube | PRH_ICU_2_tube | x | x | x | x |  |
| 10 | PRH | ICU | 3 | FH1 | Visor | PRH_ICU_3_visor | x | x | x | x |  |
|  |  |  |  |  | Tube | PRH_ICU_3_tube | x | x | x | x |  |
| 11 | PRH | ICU | 4 | FH2 | Visor | PRH_ICU_4_visor | x | x | x | x |  |
|  |  |  |  |  | Tube | PRH_ICU_4_tube | x | x | x | x |  |
| 12 | PRH | ICU | 5 | FH1 | Visor | PRH_ICU_5_visor | x | x | x | NSEM | Cracked visor |
|  |  |  |  |  | Tube | PRH_ICU_5_tube | x | x | x | x |  |
| 13 | PRH | Theatre | 1 | FH1 | Visor | PRH_Theatre_1_visor | x | x | x | NSEM |  |
|  |  |  |  |  | Tube | PRH_Theatre_1_tube | x | x | x | x |  |
| 14 | RSCH | Labour Wd | 1 | FH1 | Visor | RSCH_L13_1_visor | x | x | x | x |  |
|  |  |  |  |  | Tube | RSCH_L13_1_tube | x | x | x | x |  |
| 15 | RSCH | Labour Wd | 2 | FH1 | Visor | RSCH_L13_2_visor | x | x | x | NSEM |  |
|  |  |  |  |  | Tube | RSCH_L13_2_tube | x | x | x | x |  |
| 16 | RSCH | ENT OP | 6 | FH1 | Visor | RSCH_ENT_6_visor | x | x | x | x | Track & trace |
|  |  |  |  |  | Tube | RSCH_ENT_6_tube | Kocuria rhizophilia | x | x | x |  |
| 17 | RSCH | ENT OP | 5 | FH1 | Visor | RSCH_ENT_5_visor | x | x | x | x | Track & trace |
|  |  |  |  |  | Tube | RSCH_ENT_5_tube | x | x | x | x |  |
| 18 | RSCH | ENT OP | 1 | FH1 | Visor | RSCH_ENT_1_visor | x | x | x | x | Track & trace |
|  |  |  |  |  | Tube | RSCH_ENT_1_tube | x | x | x | x |  |
| 19 | RSCH | ENT OP | 4 | FH1 | Visor | RSCH_ENT_4_visor | x | x | x | NSEM | Track & trace |
|  |  |  |  |  | Tube | RSCH_ENT_4_tube | x | x | x | x |  |
| 20 | RSCH | ENT ward | 1 | FH1 | Visor | RSCH_L8AW_1_visor | Bacillus weihenstephensis | x | x | x |  |
|  |  |  |  |  | Tube | RSCH_L8AW_1_tube | x | x | x | x |  |
| 21 | RSCH | ED | 1 | FH2 | Visor | RSCH_ED_1_visor | x | x | x | x |  |
|  |  |  |  |  | Tube | RSCH_ED_1_tube | Micrococcus luteus | x | x | x |  |
| 22 | RSCH | ICU | 1 | FH1 | Visor | RSCH_ICU_1_visor | x | x | x | x |  |
|  |  |  |  |  | Tube | RSCH_ICU_1_tube | x | x | x | x |  |
| 23 | RSCH | ICU | 2 | FH1 | Visor | RSCH_ICU_2_visor | Staphylococcus epidermidis | x | x | NSEM |  |
|  |  |  |  |  | Tube | RSCH_ICU_2_tube | x | x | x | x |  |
| 24 | RSCH | ICU | 3 | FH1 | Visor | RSCH_ICU_3_visor | x | x | x | x |  |
|  |  |  |  |  | Tube | RSCH_ICU_3_tube | x | x | x | x |  |
| 25 | RSCH | ICU | 4 | FH2 | Visor | RSCH_ICU_4_visor | Micrococcus luteus | x | x | x |  |
|  |  |  |  |  | Tube | RSCH_ICU_4_tube | Micrococcus luteus | x | x | x |  |

PRH=Princess Royal Hospital. RSCH=Royal Sussex County Hospital. FH1/FH2=model of 3M Scott PAPR Headtop. ICU=intensive care unit; ED=emergency department; NSEM=non-sporulating environmental mould; x=negative swab; ENT=ear, nose, and throat; OP=outpatient

### 3. Swabbing PAPR hoods and tubing (simulated)

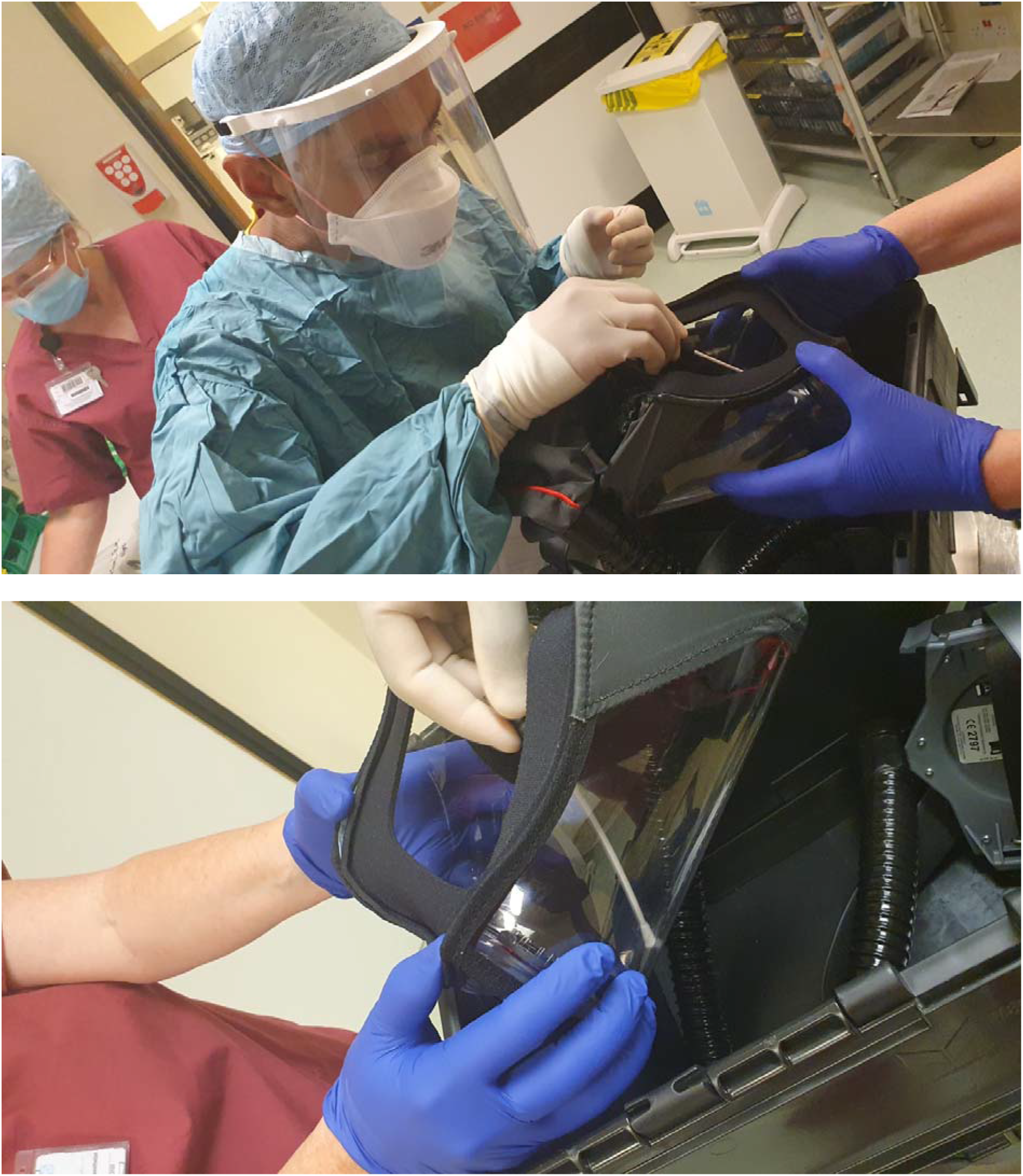

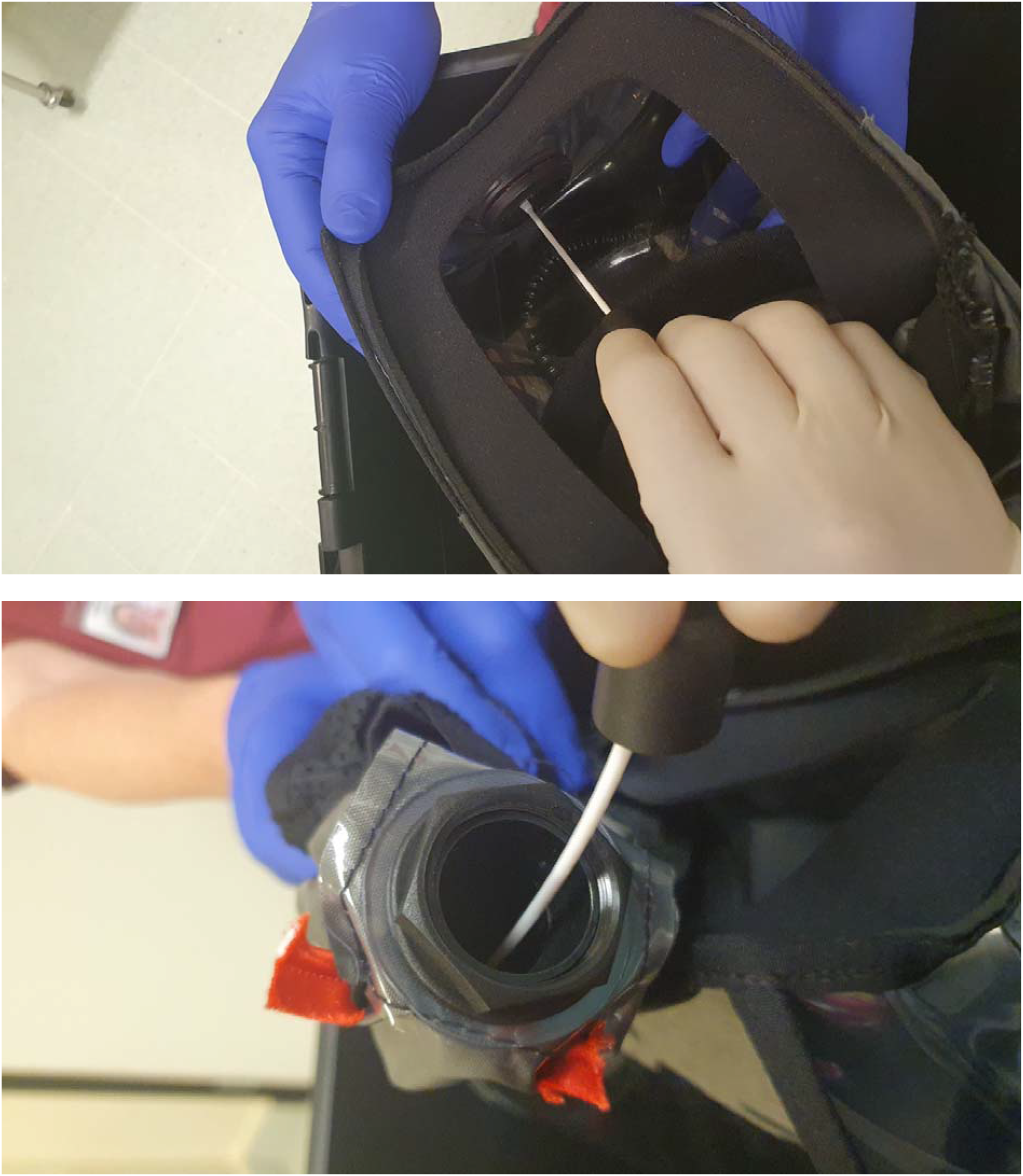
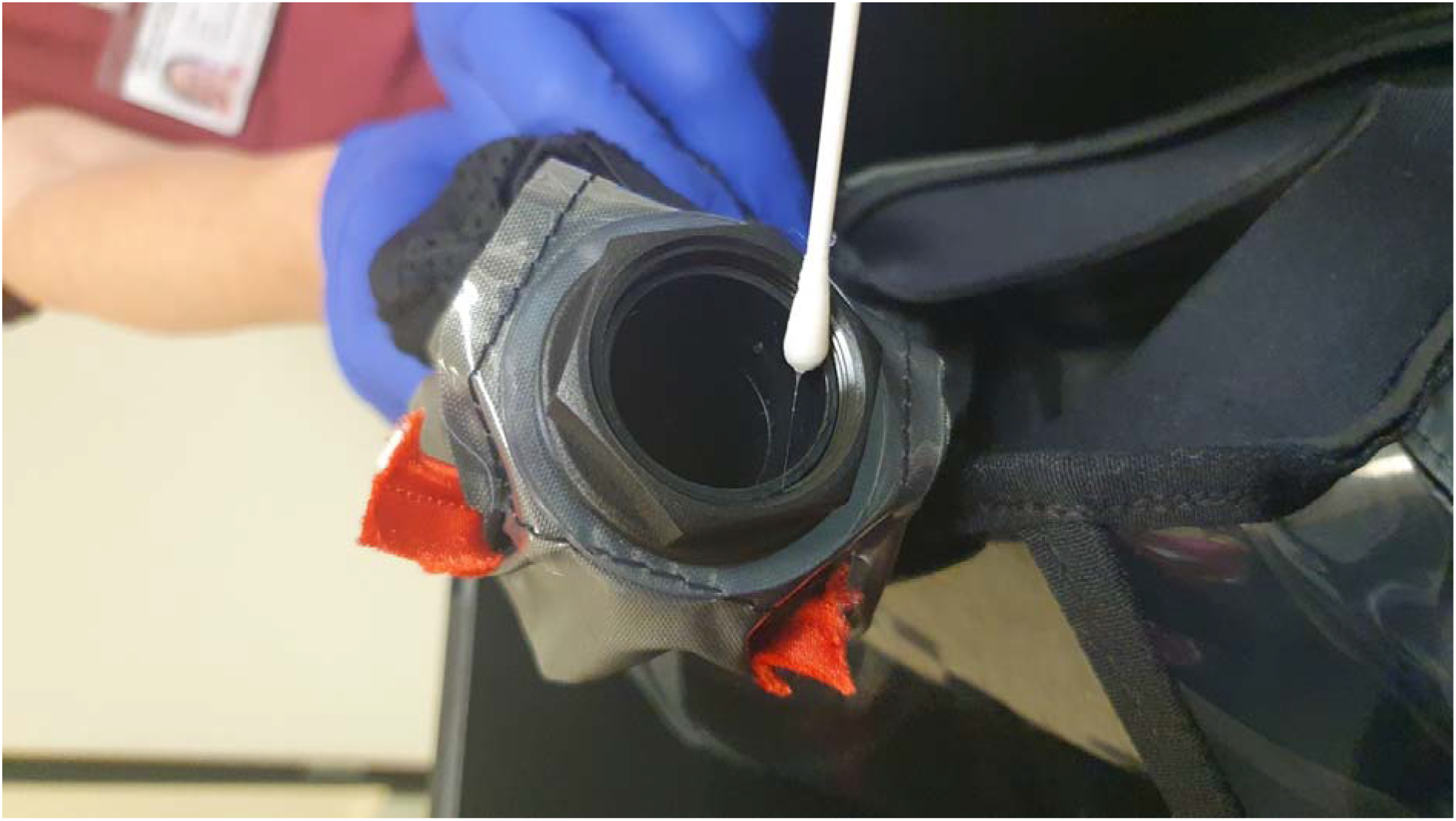

